# Twitter and Pain: An Observational Analysis of Pain-Related Tweets in Ireland

**DOI:** 10.1101/2020.04.10.20060343

**Authors:** Cormac Mullins, Robert ffrench O’Carroll, Justin Lane, Therese O’Connor

## Abstract

**Introduction:** Studies involving Twitter and chronic pain can provide highly valuable patient-generated information. The aim of this paper was to examine pain-related tweets in Ireland over a two-week period 22^nd^ June 2017-5^th^ July 2017 using pain-related keywords. We wished to identify demographic details regarding the Twitter users; most common topics discussed; sentiment analysis; and reach of tweets

**Methods:** A third-party data analytics company used 24 pain-related keywords over a 14-day period between the dates 22nd June – 5th July 2017.

**Results:** There were 941 tweets identified during the study from 715 contributors. These reached 2.62 million accounts and generated 2.88 million impressions. The most frequently occurring keywords were headache (n=321); migraine (n=147); back pain (n=123); cannabis (n=114); and chronic pain (n=85). There were 1.94 times as many tweets from females as males. The highest proportion of tweets from female users was in the fibromyalgia (83%) and migraine (60%) categories, and from males in the sciatica (35%), chronic pain (34%) and back pain (32%) categories. Cannabis-related tweets reflected mostly non-personal content (90%), with a highly positive sentiment, and the highest reach per tweet. The largest amount of advice was offered in the back pain category. Retweets were more likely to reflect a positive sentiment.

**Conclusion:** A substantial discussion of pain-related topics took place on Twitter during our study period. This provided real-time, dynamic information from individuals on discussion topics in pain medicine. This can be used to gain a greater understanding of the pain experience. As patients are increasingly acquiring healthcare information through online sources, high quality information from approved sources should be promoted on such platforms.

## INTRODUCTION

Patients are increasingly utilising internet-based forms of healthcare.^1^ These include internet-delivered interventions and health information accessed through online patient forums and social media websites, such as Facebook, YouTube, and Twitter. Internet-delivered psychological interventions in chronic pain have been demonstrated to be effective.^2^ Such models of healthcare provision are particularly attractive in areas where long waiting lists exist such as in chronic pain. Waiting times for chronic pain clinics of in excess than six months are medically unacceptable and result in significant deterioration in the clinical status of people with chronic pain (PWCP).^3, 4^

Twitter is a social media platform where users post “microblogs” or “tweets” with a maximum of 280 characters. Many individuals use Twitter for sharing thoughts and feelings, personal anecdotes, or commenting on topical items. Some users use social media to comment on medical experiences or symptoms. Infodemiology involves the use of electronic information, such as social media platforms, for the real-time study of disease characteristics and patterns.^5^ Infodemiology has been conducted on Twitter to retrieve data in a multitude of areas, such as smoking ^6^, prescription drug abuse ^7^, diet ^8^, dental pain ^9^, mood ^10^, and chronic pain.^11^

Studies involving Twitter and chronic pain can provide highly valuable patient-generated information, which is authentic and collected in real-time. This bypasses traditional problems encountered with retrospective collection methods, such as recall bias. PWCP have reported enhanced psychological, social, and cognitive outcomes from social media use.^1, 12^

The aim of this paper was to examine pain-related tweets in Ireland over a two-week period, 22^nd^ June 2017-5^th^ July 2017, using pain-related keywords. We wished to identify:

1. Demographic details regarding the Twitter users
2. Most common associated keywords/topics discussed
3. Sentiment analysis of tweets
4. Reach of pain-related tweets

## METHODS

A third-party data analytics company (Olytico, Dublin, Ireland) was used to collect data for this study. Olytico have a license to access the Twitter Firehose, which provides access to the complete stream of public messages generated on Twitter. This enabled us to search for all pain-related tweets using defined pain-related keywords. Social network analysis was limited to Twitter and geographical location restricted to Ireland. Data were collected over a 14-day period from tweets between the dates 22nd June – 5th July 2017.

24 keywords and phrases were identified. These were chronic pain; acute pain; back pain; pain clinic; Irish Pain Society; nerve pain; muscle pain; sciatica; fibromyalgia; carpal tunnel syndrome; headache; migraine; trigeminal neuralgia; slipped disc; neuropathy; myofascial; neuropathic pain; medical cannabis/cannabis/marijuana/endocannabinoids; post-herpetic neuralgia; joint pain; cancer pain; back spasm; neck pain; and Chronic Pain Ireland. Once tweets were collected, metaphorical use of pain keywords were excluded e.g. “selection headache for Gatland”. The remaining tweets represented our sample size of tweets for analysis.

Tweets were filtered by author and accounts were manually searched for on Twitter in order to identify the gender from the biographical details associated with the Twitter account. Gender was classified as “male”, “female”, or “not applicable” (N/A). “Not applicable” represented those who did not disclose their gender or represented an organisation. Individual user account names and website addresses were deleted from tweets contained within this study.

We filtered the tweets by keywords to identify the most commonly occurring keywords. The same keyword occurring twice within one tweet was counted as once. However, separate keywords occurring in the same tweet were counted in separate lists. Tweets were manually analysed to determine whether they related to oneself (“personal tweets”), or other topics (“non-personal tweets”). Non-personal tweets were further categorised into “raising awareness”, “offering advice”, and “medical research”.

Tweet content was analysed for sentiment. Tweet sentiment is an estimate of the emotion contained within the tweet. This was performed using a Twitter sentiment visualisation application, which searches for recognisable words contained within tweets in a sentiment dictionary.^13^ These words have pleasure and arousal ratings that when combined can produce an overall sentiment for the tweet. Tweets with two or more recognisable words can produce an overall sentiment rating for the tweet. Sentiment was rated on a scale of −5 to +5 of negative to positive sentiment respectively.

Dissemination of tweet content was calculated using the “potential impressions” generated. This tells us how many twitter account timelines the tweets were delivered to and gives an indication of how large the potential audience for the tweet was. Statistical analysis was performed using Statistical Package for Social Sciences (SPSS) software.^14^ For normally distributed continuous variables (e.g., sentiment analysis), the t-test was used to compare two groups (e.g., gender) and analysis of variance used to compare more than two variables (e.g., keyword categories).

## RESULTS

There were 941 tweets identified during the study from 715 contributors. This included 493 original tweets (52%), 319 retweets (34%), and 129 replies (14%). These reached 2.62 million accounts and generated 2.88 million impressions.

### Tweets by Category

The most frequently occurring keywords were headache (n=321); migraine (n=147); back pain (n=123); cannabis (n=114); chronic pain (n=85); pain “other than back pain or chronic pain” (n=57); fibromyalgia (n=34); and sciatica (n=23). Table 1 shows the most commonly occurring topics and a random sample of tweets from each topic.

**Table 1.**
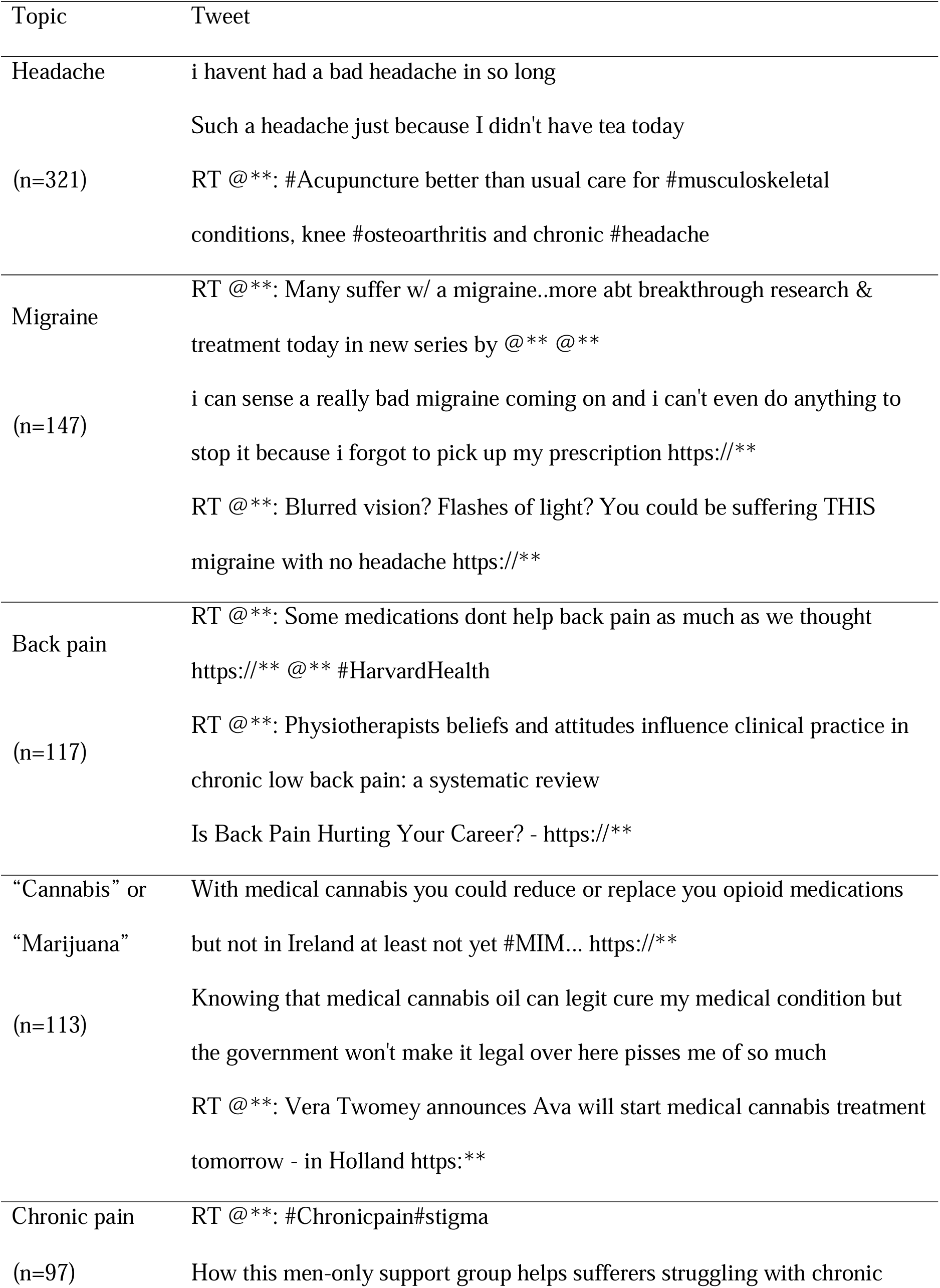
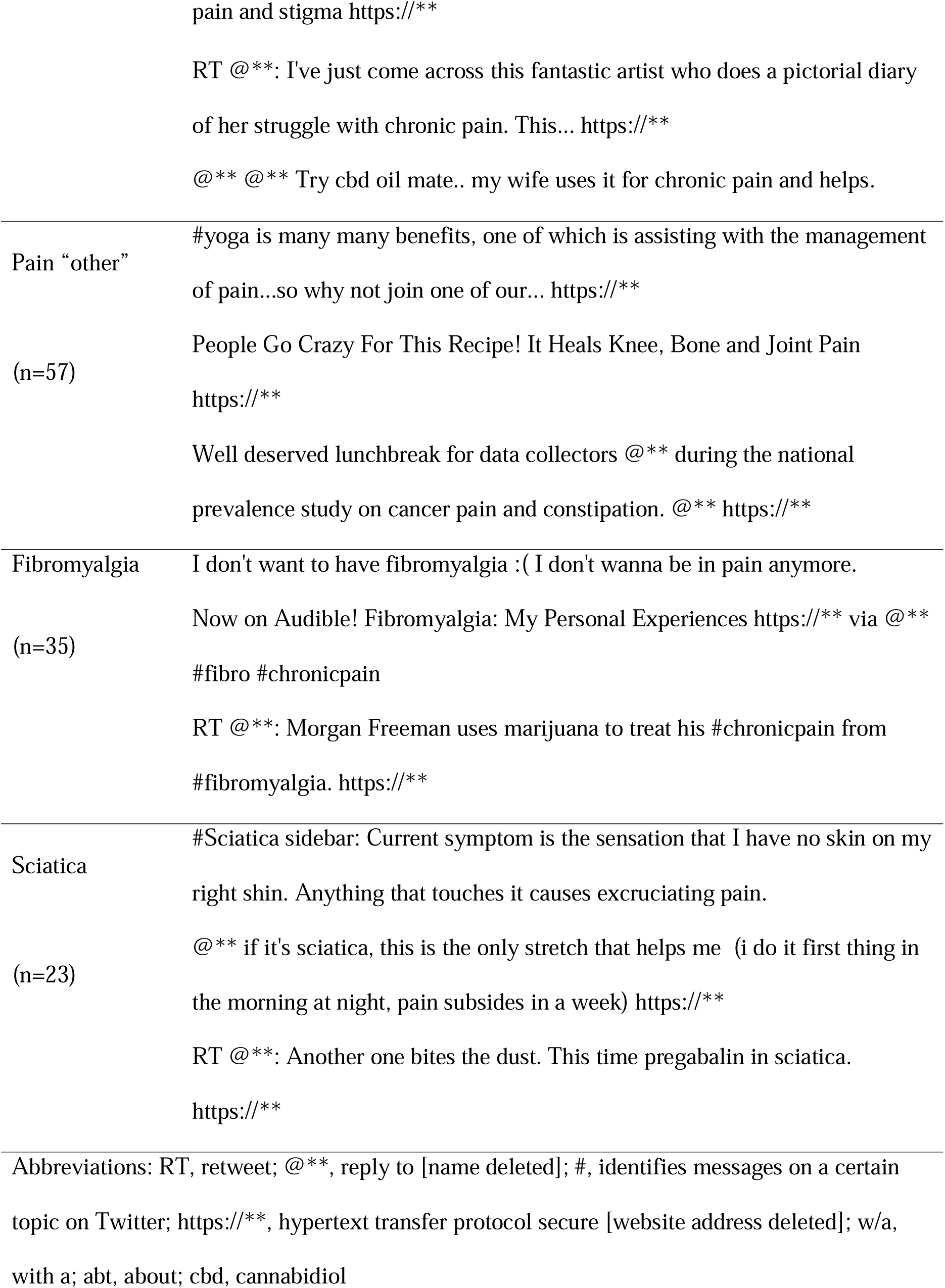
Commonly occurring topics and a random sample of tweets

### Tweets by Gender

Gender could be identified by biographical details on account information in 75% of tweets (708/941). The remaining 25% of tweets were either from accounts where gender was not disclosed, or from an organisation. There were 1.94 times as many tweets from females as males. The highest proportion of tweets from female users was in the fibromyalgia (83%) and migraine (60%) categories, while the lowest was in the cannabis category (39%) (Figure 1). The highest proportion of male tweeters was in the sciatica (35%), chronic pain (34%) and back pain (32%) category, while the lowest was in the fibromyalgia category (11%). The highest proportion of individuals who did not identify their gender or represented an organisation (“gender N/A”) was in the cannabis category (34%), and the lowest was in the fibromyalgia category (6%).

**Figure 1:**
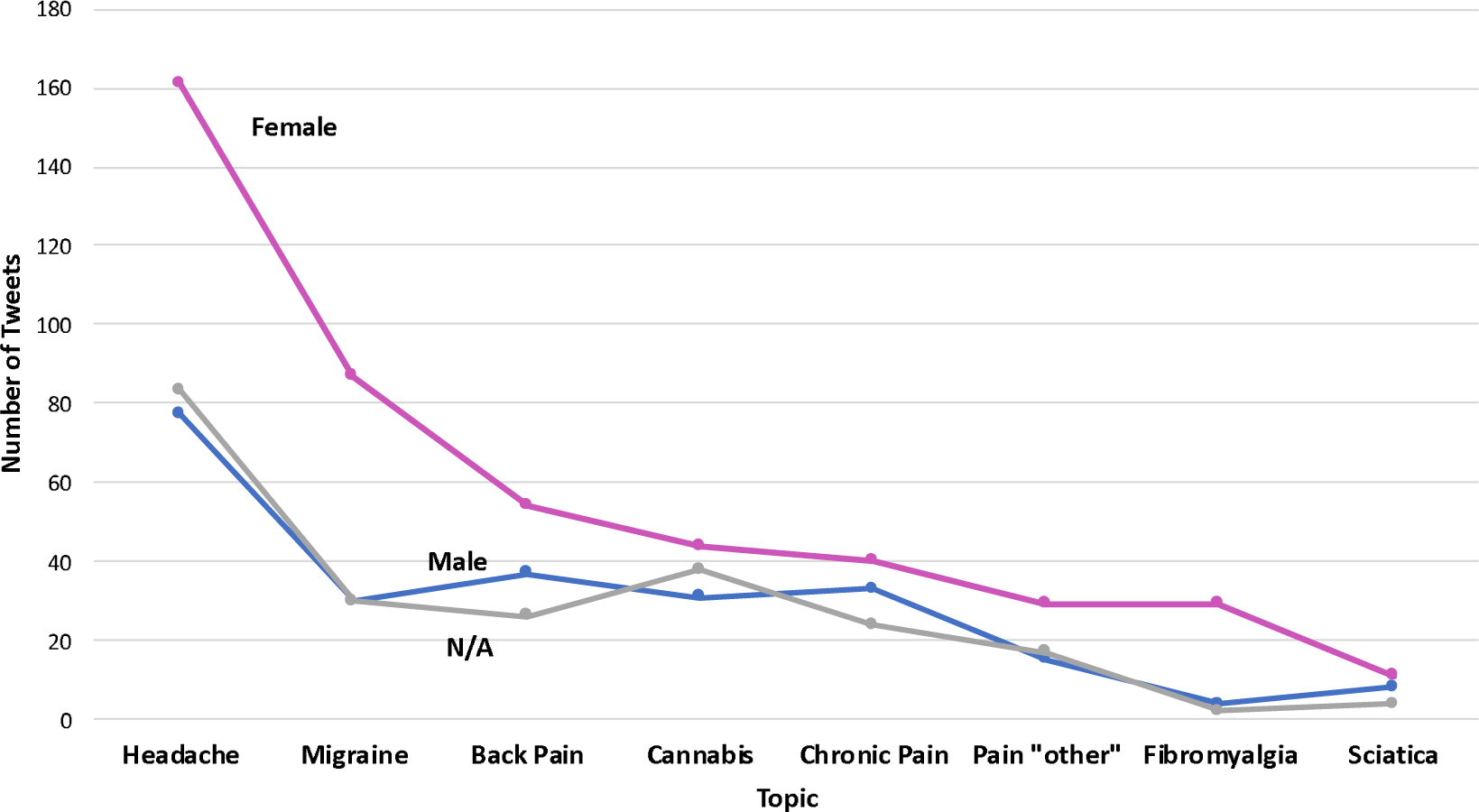
Gender Breakdown of Tweets by Topic.

### Personal and Non-Personal tweets

Personal tweets represented 52.5% of tweets (495/941). The highest proportion was in the headache (90%) and migraine (66%) categories, while the highest proportion of non-personal tweets was in the cannabis (90%) category (Figure 2). Non-personal tweets, were aimed at generating awareness (49.2%), offering advice (28.4%), or promoting medical research (18.8%) (Figure 3). Cannabis had by far the largest number of tweets aimed at generating awareness (n=99). There was a high proportion of tweets offering advice in the headache, back pain, and pain “other” categories.

**Figure 2:**
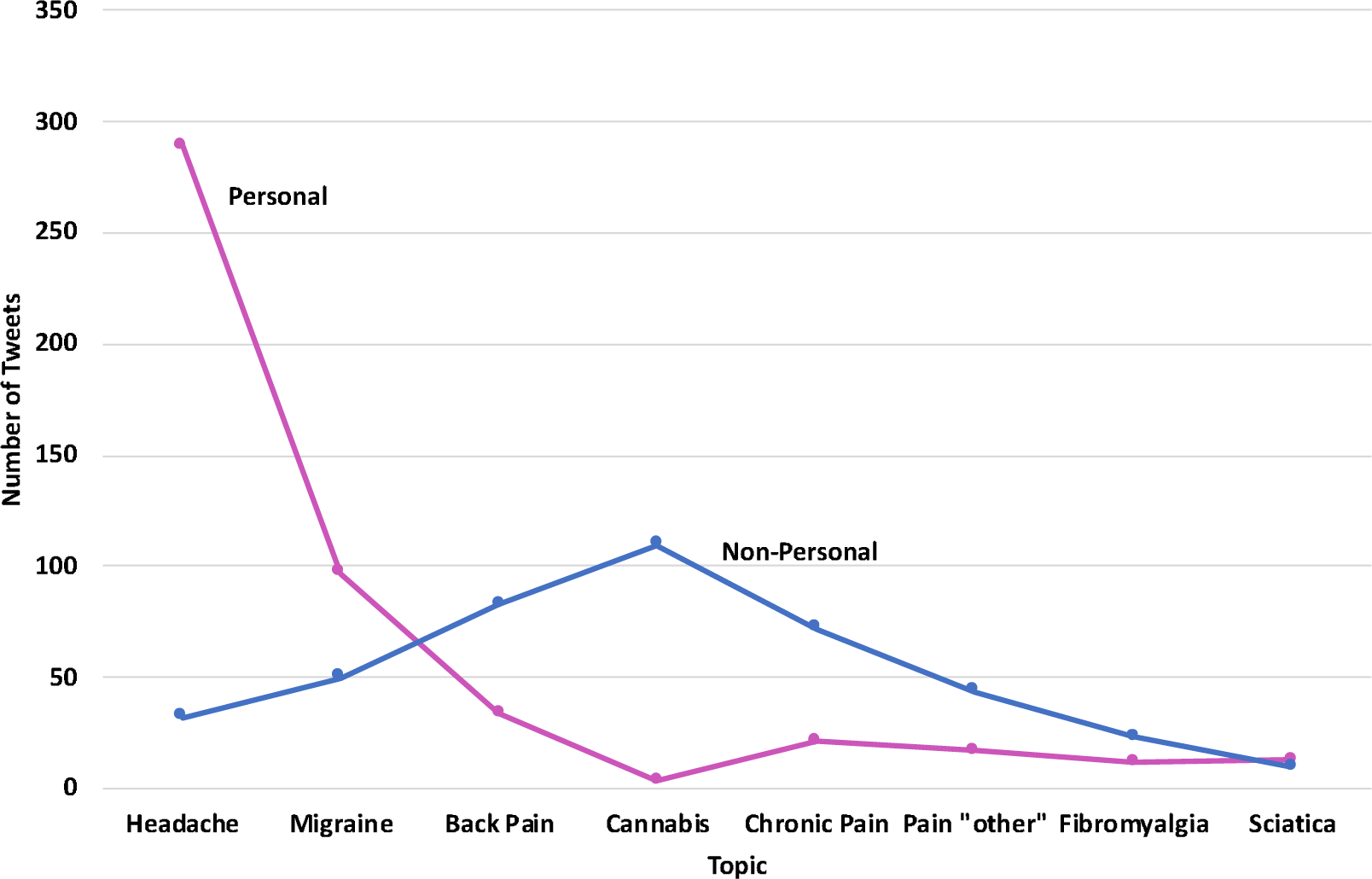
Personal or Non-Personal Tweets by Topic.

**Figure 3:**
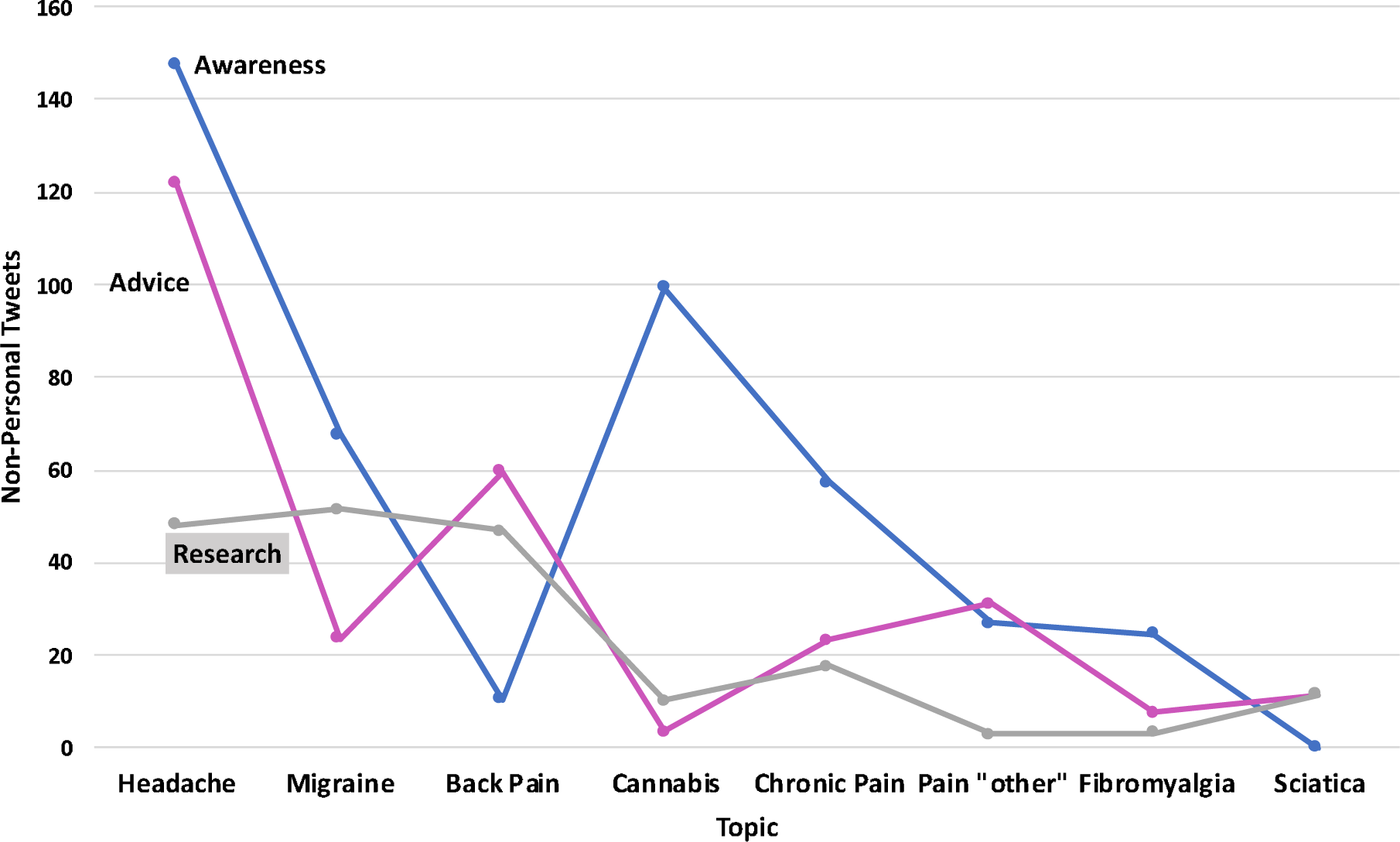
Content Breakdown of Non-Personal Tweets by Topic.

### Reach

While headache was the most common keyword, with more than double the incidence of its closest competitor, migraine still generated more impressions (618,237) compared to headache (607,473) (Figure 4). Cannabis was a close third, despite having significantly fewer keyword incidences (573,671). Back pain was the outlier with significantly fewer impressions than those beside it in the keyword table. This indicates a broad reach and a greater level of engagement for headache, migraine and cannabis on Twitter.

**Figure 4:**
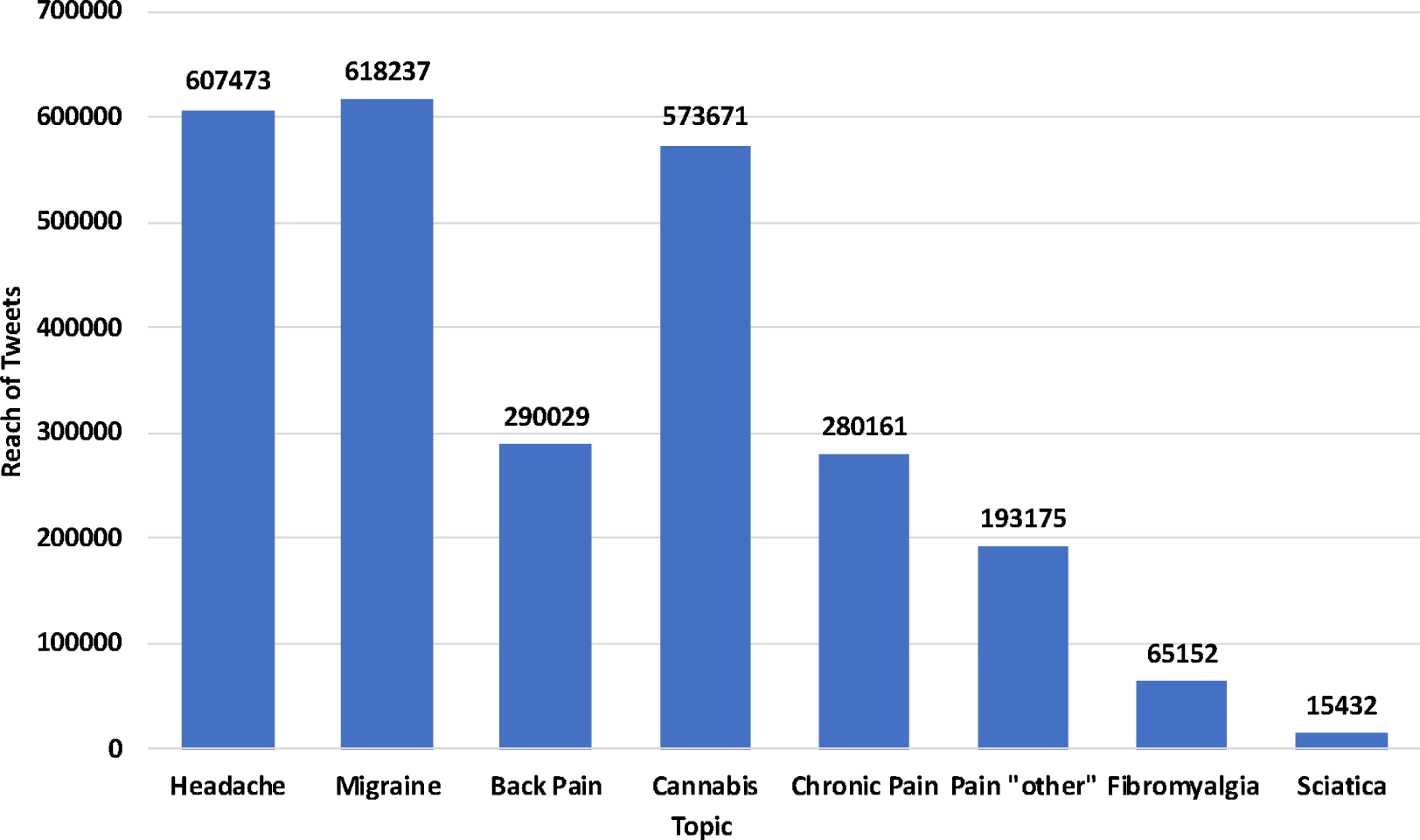
Reach of Tweets by Topic Category.

### Sentiment Analysis

There were 768 tweets with two or more recognisable terms, which allowed for an estimate of their sentiment. This represented 641 tweets and 127 retweets. The mean sentiment of all tweets was −0.17 ± 0.88; 52.5% of tweets were negative overall.

There was no sentiment disparity between the sexes (95% CI −0.1692 to 0.1662; P = 0.98). Retweets were significantly more likely to reflect a positive sentiment (95% CI 0.0817 to 0.4089; P = 0.0034). Figure 5 contains a boxplot of sentiment of tweets by topic category. The most positive sentiment was in the cannabis and fibromyalgia categories, where 83% and 68% contained an overall positive sentiment respectively.

**Figure 5:**
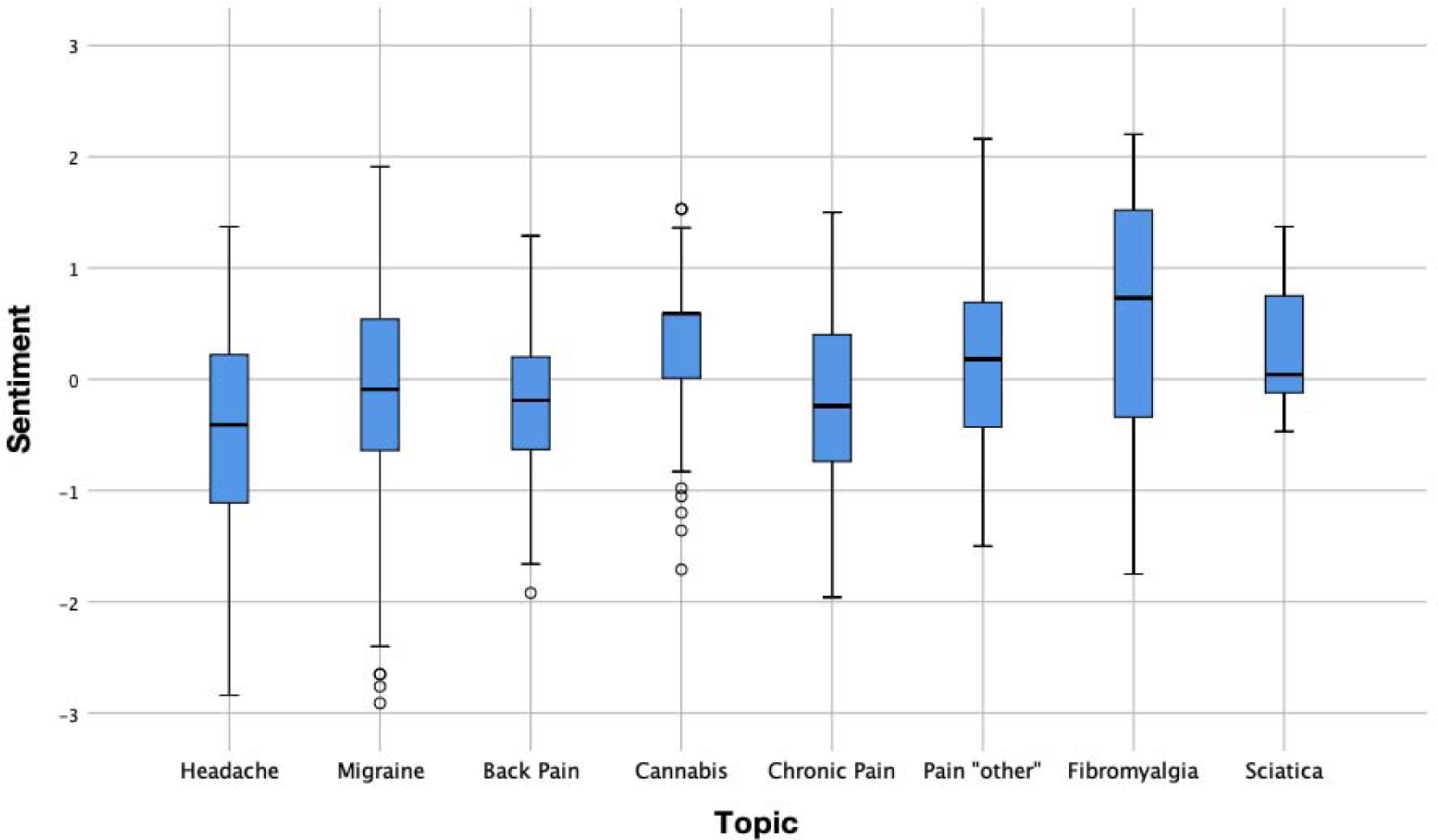
Sentiment Analysis by Topic Category.

## DISCUSSION

Twitter provides a platform for generation of real-time, unsolicited information that can provide insights into patterns and traits of many diseases in a real-world setting.^15^ Human memory is subject to recall bias; we cannot recall everything we experience and memories of experiences fade over time.^16^ Memory is selectively encoded and is influenced by factors such as emotions, duration of events, and intensity of experience.^17^ We typically remember the “peak” intensity of the pain and also how we felt at the “end” of the pain; this is described as the “peak-end effect”.^18^ This can affect recall of experiences that are repetitive over time, such as pain symptoms in chronic pain conditions. Depressed patients suffering from chronic pain are subject to cognitive distortions.^19^ This implication means that retrospective reports of pain are likely to be inaccurate. Real-time collection of social media health-related data can enhance our understanding of chronic diseases and provide an unbiased, self-expressed, and more accurate impression of pain symptoms.^20^

Social media use has been associated with improved social, psychological, and cognitive outcomes in chronic pain.^1^ Most people tend to be passive users of social media and read content rather than actively posting. Active engagement is associated with better emotional outcomes.^21^ The most common users of social media in chronic pain are well-educated females, in relationships, aged 30-60 years.^1^ In our study, 64% of those who disclosed their gender were female users. This figure rose to 70% when only tweets of personal pain symptoms were included. The highest proportion of female Twitter users was in the fibromyalgia category (83%), followed by migraine (60%), and headache (50%) (Figure 3). Fibromyalgia has been shown to be the most common underlying chronic disease among social media users with chronic pain.^1^

The most common pain-related keyword found in tweets was “headache” appearing in 34% of all tweets (n=321). Tweets containing the keyword “headache” alluded to both physical and nonphysical complaints. The words “pain” and “headache” are utilised outside of medical parlance for everyday topics and therefore can reflect a variety of themes outside of physical pain.^11^ Migraine was the most common topic outside of “headache”, present in 15.6% of tweets. While those tweeting about migraine may not have a formal diagnosis of migraine, it has been shown that those who report their headache symptoms as migraine are three times more likely to legitimately suffer from this.^22^ The majority of tweets related to headache (90.3%) and migraine (66%) were generated from patients’ report of ongoing symptoms. The majority of tweets had a predominantly negative sentiment, which is unsurprising.

### Advice on Twitter

Advice surrounding back pain and headache made up a significant proportion of these categories. Patients are increasingly sharing their personal medical experiences online; the majority of online material on scientific forums is predominantly based on opinions and personal experience, rather than on scientific results.^23^ In USA, 41% of e-patients have read other patients’ personal medical stories on an online blog or website and 40% of Americans doubt their doctor’s professional opinion when it conflicts with online information from social media websites.^24^ Users of health services, particularly those who are on long waiting lists to access health services can be vulnerable to misleading information. As healthcare professionals, we should advise caution on the content of healthcare information that is promoted on social media platforms. Patients accessing material online may not have the expertise to filter material that is not scientifically rigorous. This can have a negative impact, as demonstrated by the adverse effect of online parents’ discussion forums on the uptake of childhood vaccination.^25^ Furthermore, the quality of retrieved material is dependent on the search terminology.^26^ As patients actively search for online material themselves, material may not be searched for in a wholly objective manner and may be subject to confirmation bias.^27^ A systematic review found that while high quality material produced by government and professional organisations is accessible on YouTube, there also exists poor quality information, primarily anecdotal, which online users have a high likelihood of coming into contact with.^26^ In light of this, it is important that high-quality health information from approved sources be promoted on these platforms to ensure vulnerable individuals are not exploited by misleading advice or information.

### Medical Cannabis and Twitter

“Cannabis” tweets had a unique set of characteristics compared to the other categories. They were mostly “non-personal” (97%), awareness-promoting (88%), with a highly positive sentiment (83%), and the greatest reach per tweet. They also had the highest proportion of users who represented an organisation. The reasons these figures could differ are manifold. First of all, cannabis use is highly restricted in the Republic of Ireland and is not available for chronic pain conditions according to the Health Products Regulatory Agency (HPRA) recommendations on the use of cannabis products for medical use in Ireland.^28^ This is at odds with the European Pain Federation (EFIC) position paper on the use of cannabis products in chronic pain conditions.^29^ It also differs from the other categories in our study, in that it is a form of treatment rather than a symptom or a diagnosis. Therefore, individuals are unlikely to be tweeting about their use of medical cannabis in Ireland for chronic pain conditions.

Medical cannabis is evidently is a controversial topic, as demonstrated by the conflicting statements from the HPRA and EFIC. Accordingly, this topic generated the highest number of impressions per tweet. While sentiment analysis was mostly positive for cannabis-related tweets, it is difficult to comment on how this data reflects public opinion without comparing it to other data points such as survey data of the public on this topic. It is quite likely that this positive attitude reflected on social media to cannabis represents a vocal minority of the population who are eager to champion a cause that is important to them.^30^ Often, certain views can appear more mainstream than they actually are due to greater activity of a vocal minority over a silent majority.^30^ It is also possible that these views being promoted on social media could represent an underlying specific agenda. While people frequently utilise social media to advocate for certain causes, a vocal minority can distort this discourse.

Promotional activities have begun to play a large role in social media discourse. The Cambridge Analytica hearings demonstrated that data can be used to strategically to manipulate and to deceive public opinion.^31^ There appears to be a disparity between public perception of efficacy and safety of cannabis-related products and the reality.^28^ The volume of posts and information present on social media platforms can it difficult to broadcast scientifically rigorous data. Governmental attempts to direct such discourse may do more harm than good.^32^ Public life has evolved to incorporate multiple complex and interwoven public domains including Twitter and other social media platform and these can have a significant effect on public opinion.^33^ Further work needs to be done on ascertaining whether social media data on these issues actually reflects an opinion that is held by the wider society.

### Concerns about social media

Other concerns exist regarding social media use among patients including regulation of privacy of data, doctor-patient confidentiality, and loss of control over information provided to patient.^34^ Some of these concerns can be addressed and require engagement on the part of physicians. It is essential that healthcare professionals provide leadership in this area by facilitating access to reliable, scientifically valid online information, and through the development of best practice guidelines for accessing information for patients. The development of social media guidelines for healthcare professionals and incorporation into medical student education is essential to ensure optimal integration of novel media platforms into modern medicine.^35^ Novel approaches are needed to support a symbiosis between patients and physicians on social media.^36^ Physicians and public health specialists need to adapt to the new ways in which patients consume information.^37^ Many patients are rating their healthcare experiences online. Insights gained from social media posting from patient populations can be utilised by physicians to gain a valuable perspective on their patients’ worldview. Adapting to patients’ behaviour can result in greater patient engagement, patient satisfaction, quality of care, and improved health outcomes.^38^ This can enrich the doctor-patient therapeutic relationship.^39^

### Healthcare professionals and Twitter

Healthcare professional are increasingly using Twitter for professional and academic reasons. This has taken the form of virtual journal clubs ^40^, Twitter-enhanced medical conferences ^41^, distribution of academic and research material ^42^, and advertising of professional services. Twitter has been used to conduct research and highly tweeted research is more likely to be cited.^42^ Virtual journal clubs involve online interactive discussions on journal papers. This provides a novel take on reading and critiquing journal articles and is not constrained by the usual limitations of a live journal club meeting. Twitter-enhanced medical conferences increase reach of material to non-attendees and the general public. The use of a pre-conference Twitter campaign has been demonstrated to increase Twitter engagement at medical conferences.^43^ Social media has also been used successfully to promote educational videos to the general public on chronic pain related material.^44^

### Limitations

This study was conducted over a two-week study period. Public discourse on social media can change rapidly and is dictated by world events. Therefore, our findings may be distorted by events that were topical during this timeframe. Ideally, this study should be replicated over a longer timeframe to establish the consistency of our findings. Furthermore, the geographical location of tweets was restricted to Ireland. It is unclear whether the pattern of Twitter use described here is comparable internationally.

Assessment of sentiment, particularly within the context of a 280-character limit and which may contain abbreviations, slang, and “smileys”, raises questions over the validity and reliability of such data. Twitter sentiment analysis has been used successfully in other studies, but assessing a change in sentiment may be difficult and can be biased towards a negative change.^45, 46^ Comparison of this information with other data points, such as survey data should ideally be undertaken.

## CONCLUSION

A substantial discussion of pain-related topics took place on Twitter during our study period. This provided real-time, dynamic information from individuals on discussion topics in pain medicine. This can be used to gain a greater understanding of the pain experience. As patients are increasingly acquiring healthcare information through online sources, high quality information from approved sources needs to be promoted on such platforms. New ways of thinking are required to ensure satisfactory incorporation of social media into mainstream medicine.

## Data Availability

The authors confirm that the data supporting the findings of this study are available within the article.

## Acknowledgments

We would like to acknowledge Dr. Christopher Healey, North Carolina State University for use of North Carolina University sentiment analysis software. We would also like to acknowledge Olytico, Dublin, Ireland for conducting the Twitter search.

## REFERENCES

1. Merolli M, Gray K, Martin-Sanchez F, Lopez-Campos G. Patient-reported outcomes and therapeutic affordances of social media: findings from a global online survey of people with chronic pain. J Med Internet Res. 2015;17(1):e20.

2. Buhrman M, Skoglund A, Husell J, Bergstrom K, Gordh T, Hursti T, et al. Guided internet-delivered acceptance and commitment therapy for chronic pain patients: a randomized controlled trial. Behav Res Ther. 2013;51(6):307–15.

3. Lynch ME, Campbell F, Clark AJ, Dunbar MJ, Goldstein D, Peng P, et al. A systematic review of the effect of waiting for treatment for chronic pain. Pain. 2008;136(1-2):97–116.

4. Dear BF, Titov N, Perry KN, Johnston L, Wootton BM, Terides MD, et al. The Pain Course: a randomised controlled trial of a clinician-guided Internet-delivered cognitive behaviour therapy program for managing chronic pain and emotional well-being. Pain. 2013;154(6):942–50.

5. Eysenbach G. Infodemiology and infoveillance: framework for an emerging set of public health informatics methods to analyze search, communication and publication behavior on the Internet. J Med Internet Res. 2009;11(1):e11.

6. Myslin M, Zhu SH, Chapman W, Conway M. Using twitter to examine smoking behavior and perceptions of emerging tobacco products. J Med Internet Res. 2013;15(8):e174.

7. Hanson CL, Cannon B, Burton S, Giraud-Carrier C. An exploration of social circles and prescription drug abuse through Twitter. J Med Internet Res. 2013;15(9):e189.

8. Hingle M, Yoon D, Fowler J, Kobourov S, Schneider ML, Falk D, et al. Collection and visualization of dietary behavior and reasons for eating using Twitter. J Med Internet Res. 2013;15(6):e125.

9. Heaivilin N, Gerbert B, Page JE, Gibbs JL. Public health surveillance of dental pain via Twitter. J Dent Res. 2011;90(9):1047–51.

10. Golder SA, Macy MW. Diurnal and seasonal mood vary with work, sleep, and daylength across diverse cultures. Science. 2011;333(6051):1878–81.

11. Tighe PJ, Goldsmith RC, Gravenstein M, Bernard HR, Fillingim RB. The painful tweet: text, sentiment, and community structure analyses of tweets pertaining to pain. J Med Internet Res. 2015;17(4):e84.

12. Tsai S, Crawford E, Strong J. Seeking virtual social support through blogging: A content analysis of published blog posts written by people with chronic pain. Digit Health. 2018;4:2055207618772669.

13. Computer Science Department NCSU. Twitter Sentiment Visualisation 2020 [

14. Corp. I. IBM SPSS Statistics for Mac, Version 26.0. Armonk, NY: IBM Corp. Released 2019.

15. Stone AA, Broderick JE. Real-time data collection for pain: appraisal and current status. Pain Med. 2007;8 Suppl 3:S85–93.

16. Kikuchi H, Yoshiuchi K, Miyasaka N, Ohashi K, Yamamoto Y, Kumano H, et al. Reliability of recalled self-report on headache intensity: investigation using ecological momentary assessment technique. Cephalalgia. 2006;26(11):1335–43.

17. Walentynowicz M, Bogaerts K, Van Diest I, Raes F, Van den Bergh O. Was it so bad? The role of retrospective memory in symptom reporting. Health Psychol. 2015;34(12):1166–74.

18. Redelmeier DA, Kahneman D. Patients’ memories of painful medical treatments: real-time and retrospective evaluations of two minimally invasive procedures. Pain. 1996;66(1):3–8.

19. Smith TW, O’Keeffe JL, Christensen AJ. Cognitive distortion and depression in chronic pain: association with diagnosed disorders. J Consult Clin Psychol. 1994;62(1):195–8.

20. Gendreau M, Hufford MR, Stone AA. Measuring clinical pain in chronic widespread pain: selected methodological issues. Best Pract Res Clin Rheumatol. 2003;17(4):575–92.

21. Setoyama Y, Yamazaki Y, Namayama K. Benefits of peer support in online Japanese breast cancer communities: differences between lurkers and posters. J Med Internet Res. 2011;13(4):e122.

22. Lipton RB, Stewart WF, Liberman JN. Self-awareness of migraine: interpreting the labels that headache sufferers apply to their headaches. Neurology. 2002;58(9 Suppl 6):S21–6.

23. Sudau F, Friede T, Grabowski J, Koschack J, Makedonski P, Himmel W. Sources of information and behavioral patterns in online health forums: observational study. J Med Internet Res. 2014;16(1):e10.

24. Sadah SA, Shahbazi M, Wiley MT, Hristidis V. Demographic-Based Content Analysis of Web-Based Health-Related Social Media. J Med Internet Res. 2016;18(6):e148.

25. Brunson EK. The impact of social networks on parents’ vaccination decisions. Pediatrics. 2013;131(5):e1397–404.

26. Madathil KC, Rivera-Rodriguez AJ, Greenstein JS, Gramopadhye AK. Healthcare information on YouTube: A systematic review. Health Informatics J. 2015;21(3):173–94.

27. Nickerson RS. Confirmation bias: A ubiquitous phenomenon in many guises. Review of general psychology. 1998;2(2):175–220.

28. Agency HPR. Cannabis for medical use. A scientific review. 2017.

29. Hauser W, Finn DP, Kalso E, Krcevski-Skvarc N, Kress HG, Morlion B, et al. European Pain Federation (EFIC) position paper on appropriate use of cannabis-based medicines and medical cannabis for chronic pain management. Eur J Pain. 2018;22(9):1547–64.

30. Mustafaraj E, Finn S, Whitlock C, Metaxas PT, editors. Vocal minority versus silent majority: Discovering the opionions of the long tail. Privacy, Security, Risk and Trust (PASSAT) and 2011 IEEE Third Inernational Conference on Social Computing (SocialCom), 2011 IEEE Third International Conference on; 2011: IEEE.

31. Isaak J, Hanna MJ. User data privacy: Facebook, Cambridge Analytica, and privacy protection. Computer. 2018;51(8):56–9.

32. Shirky C. The political power of social media: Technology, the public sphere, and political change. Foreign affairs. 2011:28–41.

33. Keane J. Structural transformations of the public sphere. The Media, Journalism and Democracy: Routledge; 2018. p. 53–74.

34. Hughes B, Joshi I, Wareham J. Health 2.0 and Medicine 2.0: tensions and controversies in the field. J Med Internet Res. 2008;10(3):e23.

35. Knight E, Werstine RJ, Rasmussen-Pennington DM, Fitzsimmons D, Petrella RJ. Physical therapy 2.0: leveraging social media to engage patients in rehabilitation and health promotion. Phys Ther. 2015;95(3):389–96.

36. Rozenblum R, Greaves F, Bates DW. The role of social media around patient experience and engagement. BMJ Qual Saf. 2017;26(10):845–8.

37. Suman A, Bostick GP, Schopflocher D, Russell AS, Ferrari R, Battie MC, et al. Long-term evaluation of a Canadian back pain mass media campaign. Eur Spine J. 2017;26(9):2467–74.

38. Miller KL. Patient centered care: A path to better health outcomes through engagement and activation. NeuroRehabilitation. 2016;39(4):465–70.

39. Modica RF, Lomax KG, Batzel P, Cassanas A. Impact of systemic juvenile idiopathic arthritis/Still’s disease on adolescents as evidenced through social media posts. Open Access Rheumatol. 2018;10:73–81.

40. Thangasamy IA, Leveridge M, Davies BJ, Finelli A, Stork B, Woo HH. International urology journal club via Twitter: 12-month experience. European urology. 2014;66(1):112–7.

41. Pemmaraju N, Mesa RA, Majhail NS, Thompson MA, editors. The use and impact of Twitter at medical conferences: best practices and Twitter etiquette. Seminars in hematology; 2017: Elsevier.

42. Peoples BK, Midway SR, Sackett D, Lynch A, Cooney PB. Twitter Predicts Citation Rates of Ecological Research. PLoS One. 2016;11(11):e0166570.

43. Schwenk ES, Jaremko KM, Gupta RK, Udani AD, McCartney CJL, Snively A, et al. Upgrading a Social Media Strategy to Increase Twitter Engagement During the Spring Annual Meeting of the American Society of Regional Anesthesia and Pain Medicine. Reg Anesth Pain Med. 2017;42(3):283–8.

44. White R, Hayes C, White S, Hodson FJ. Using social media to challenge unwarranted clinical variation in the treatment of chronic noncancer pain: the “Brainman” story. J Pain Res. 2016;9:701–9.

45. Bollen J, Mao H, Zeng X. Twitter mood predicts the stock market. Journal of computational science. 2011;2(1):1–8.

46. Thelwall M, Buckley K, Paltoglou G. Sentiment in Twitter events. Journal of the American Society for Information Science and Technology. 2011;62(2):406–18.

